# Vaccination or restriction?: COVID-19 vaccine hesitancy and vaccine passports

**DOI:** 10.1101/2021.09.15.21263559

**Authors:** Shohei Okamoto, Kazuki Kamimura, Kohei Komamura

## Abstract

**Objectives:** While the development of vaccines against the novel coronavirus (COVID-19) brought the hope of establishing herd immunity, which might help end the global pandemic, vaccine hesitancy can hinder the progress towards herd immunity. In this study, we assess the determinants of vaccine hesitancy, reasons for hesitation, and effectiveness of vaccine passports in relaxing public health restrictions.

**Methods:** Through an online survey that includes a conjoint experiment of a demographically representative sample of 5,000 Japanese adults aged 20–74, we assess the determinants of vaccine hesitancy, reasons for hesitation, and effectiveness of hypothetical vaccine passports.

**Results:** We found that about 30% of respondents did not intend to vaccinate or have not yet decided, with major reasons for vaccine hesitancy being related to concerns about the safety and side effects of the vaccine. In line with previous findings, younger age, lower socioeconomic status, and psychological factors such as weaker COVID-19 fear were associated with vaccine hesitancy. The easing of public health restrictions such as travel, wearing face masks, and dining out at night was associated with an increase in vaccine acceptance by 4–10%.

**Conclusion:** Vaccine hesitancy can be reduced by mitigating the concerns about vaccine safety and side effects, as well as by relaxing public health restrictions. However, the feasibility of vaccine passports needs to be sufficiently assessed, taking the ethical issues of passports and the public health impacts of the relaxation of restrictions into careful consideration.

**Strengths and limitations of this study:**

□ This study includes timely data on COVID-19 vaccine hesitancy, obtained from a demographically representative sample of 5,000 Japanese adults.
□ A conjoint experiment allows assessing the effectiveness of easing public health restrictions on vaccine acceptance.
□ Actual behaviour may diverge from the survey responses or fluctuate due to the pandemic situation and the timing of the survey.
□ Results may not be applicable in other countries, since the pandemic situation, government responses to the pandemic, and reasons for vaccine hesitancy can vary across countries.

## Introduction

After a period when nations have managed to curb the spread of the new coronavirus disease (COVID-19), mainly by non-pharmaceutical interventions such as containment and closure policies, the development of COVID-19 vaccines brought hope that the pandemic may end soon. Although the degree and duration of vaccine efficacy as well as the efficacy against new virus variants remain unconfirmed, widespread vaccination can contribute to establishing herd immunity against COVID-19. While the required proportions of individuals with immunity could vary by country (e.g. due to demographic structure and frequency of human contact), it is estimated that approximately 70% of the population needs immunity to achieve herd immunity against COVID-19, which would require more than 30 million deaths worldwide due to natural infection [1]. Therefore, global vaccination is a necessary step to end the pandemic.

However, the vaccine hesitancy, defined as a ‘delay in acceptance or refusal of vaccines despite availability of vaccine services’ by a working group advising the World Health Organization [2], can hinder achieving herd immunity. The findings of systematic reviews and meta-analyses suggest that the vaccine acceptance rates are around 60–75% but show large discrepancies across regions, months of study, and whether an answer of ‘unsure’ is available to survey respondents [3, 4]. Together with the global disparities in vaccine availability for COVID-19, full vaccination rates are considerably low, and only about a third of the world population had received at least one dose of a vaccine against COVID-19 by August 2021 [5]. While this low vaccine uptake may be due to many reasons, including factors other than individual preferences, such as system failures, it is important to identify why people are reluctant to vaccinate to establish herd immunity by avoiding preventable vaccine hesitancy.

Whether an individual accepts vaccination is a consequence of a complex decision-making process, which occurs on the continuum between complete acceptance and refusal [2]. The above-mentioned working group developed the ‘three Cs vaccine hesitancy model’, which comprises confidence, complacency, and convenience, indicating that historic, socio-cultural, environmental, health system/institutional, economic or political factors, as well as personal perception and vaccine/vaccination characteristics matter towards vaccine hesitancy. Moreover, from a utilitarian perspective, voluntary vaccinations can deviate from the social optimum due to the positive externalities of vaccinated individuals; hence, Pigouvian subsidies, external regulations, or strategies to improve vaccine awareness is needed, depending on the nature of vaccine-preventable diseases and vaccines [6, 7].

Therefore, it is important to understand the reasons of those who tend not to accept a COVID-19 vaccine, in addition to strategies to raise the vaccination rates. In the following section, we review the literature on the determinants of COVID-19 vaccine hesitancy.

### Literature on COVID-19 vaccine hesitancy

#### (1) Socio-demographic factors

Given the concerns of the increasing hesitancy towards COVID-19 vaccination, many empirical studies have hitherto assessed factors associated with COVID-19 vaccine hesitancy [3, 8-11]. These studies suggest that many empirical papers find that older people compared to their younger counterparts and men compared to women are more likely to accept a COVID-19 vaccine. Older people are susceptible to the disease and while men and women can decline vaccination for different reasons, the differences in perceived risks, efficacies, and knowledge may mediate these gender differences [11]. In addition to age and gender, educational attainment is identified as the most frequent predictor, with higher acceptance among people with higher education levels [3]. While highly educated individuals can be vaccine-hesitant because of the influence of social groups and other authorities, education may play an important role in understanding disease severity and vaccination benefits [11].

#### (2) Psychological and behavioural factors

Vaccination is a consequence of one’s utility maximisation, considering costs and benefits. Based on the health belief model, by modifying socio-demographic factors, individuals can decide whether to vaccinate as a reflection of their personal beliefs about a disease and its preventive measures, such as susceptibility, severity, benefits, barriers, and self-efficacy [12, 13]. A systematic review identifies that vaccine acceptance is higher among those with greater perceived risk, threat, vulnerability, and susceptibility to infection [8, 9]. Furthermore, the beliefs about the vaccine are predictors of vaccine hesitancy, including mistrust in its safety or efficacy and conspiracy beliefs, which can be induced by a low health literacy and negative information in the media [8].

Together with one’s perceptions regarding vaccines and infection, individual preferences matter for health-related decision-making, including vaccination [14, 15]. Time preference affects one’s vaccination intentions because individuals will benefit from the vaccination in the future, despite having to bear its present costs. Therefore, those discounted future benefits would lead them to decide not to vaccinate. Moreover, the attitudes toward risks are attributed to vaccination decision-making, that is, risk-averse individuals would feel conflicted between the risk of infection without vaccination and the vaccines’ side effects. This would explain why younger people tend to be vaccine-hesitant, considering they are less likely to develop symptoms than their older counterparts [16] and have more frequent side effects [17]. In addition to individuals’ attributes and beliefs, vaccine characteristics are important determinants of vaccination intention, being highly relevant to individuals’ perceptions and preference for vaccines. In particular, individuals prefer vaccines with higher efficacy, longer duration of disease protection, and safety (i.e. none or few adverse effects) [8, 18].

### Vaccination campaigns

To increase vaccination rates, considering the determinants of vaccine hesitancy discussed above and vaccine characteristics, potential strategies would include removing the (mis)perceived effectiveness and risks of vaccination and infection, minimising the costs associated with vaccination (i.e. out-of-pocket payments and opportunity cost), and increasing the benefits of vaccination by providing various incentives. In fact, several approaches, such as communication, financial and non-financial incentives, and reminder-recall interventions, have been adopted and evaluated so far [19, 20].

Vaccination campaign frameworks have also been adopted to increase the COVID-19 vaccination uptake. Aiming at a better understanding of the population for COVID-19 vaccines, public organisations disseminate information about the efficacy and safety of vaccines [21, 22]. Additionally, a study suggests that emphasising the benefits of vaccination and inducing feelings of vaccine ownership are useful [23, 24], thus suggesting the importance of information campaigns. In some countries, the convenience of vaccination locations is enhanced by providing these services within the areas of citizens’ daily lives, such as train stations and supermarkets [25, 26]. Furthermore, the incentives toward vaccination attract the attention of some policymakers [27] although their effectiveness remains inconclusive and may depend on how incentives are given [28, 29].

With remaining ethical concerns, vaccine passports, which fully or partially exempt vaccinated individuals from public health restrictions [30, 31], are considered in many regions. While only a limited number of related studies are available, the freedom allowed by vaccine passports can affect vaccine acceptance and preference [29, 32]. Despite the concern about ‘breakthrough infections’, it would be worth considering the applicability of vaccine passports if and only if the passports largely contribute to achieving herd immunity against COVID-19 by increasing vaccine acceptance.

### Contributions of this study

Previous studies have documented the determinants of vaccine hesitancy by analysing the association of socio-demographic, psychological, and vaccine characteristics with vaccine intentions. Meanwhile, the evidence on how to increase COVID-19 vaccine acceptance remains scarce. In alignment with the policies in several regions, the evidence on communication strategies and the incentives for reducing vaccine hesitancy have gained increasing attention, as discussed above, while the effectiveness of vaccine passports in raising vaccine acceptance has been limited. While countries such as Israel, France, and Italy attempt to utilise vaccine passports, other countries may also consider similar schemes to return to a ‘normal life’. If so, it is immensely important to accelerate herd immunity by reducing avoidable vaccine hesitancy, to which the benefits of the vaccine passports may contribute.

Therefore, we assess the effectiveness of easing public health restrictions after vaccination by vaccine passports based on our analysis of a conjoint experiment. By decomposing the freedom factors allowed by the passport based on government regulations, we first evaluate an effective type of relaxation of public health restrictions to increase vaccine acceptance, which would be useful for health policymakers to design vaccine passports and deliver attractive information on the benefits of vaccination for vaccine-hesitant individuals.

## Methods

### Data

The data come from a demographically representative sample of 5,000 Japanese adults aged 20–74, which was conducted online during 21–23 July 2021. Survey respondents were recruited from registered panels of Cross Marketing Inc. To ensure that the survey is nationally representative regarding respondents’ age and gender, we recruited respondents using quota sampling for each of the 14 age groups of the 2015 population census (i.e. age categories of 20s, 30s, 40s, 50s, 60s, and early 70s by gender).

While we did not use regional quotas to recruit respondents, we addressed the potential non-representativeness arising from this by using weights for all analyses and estimated by population structures in each region. Specifically, we used eight categories for residential areas (i.e. Hokkaido, Tohoku, and Kanto, except for the Tokyo Metropolitan Area, Tokyo Metropolitan Area, Chubu, Kinki, Chugoku, Shikoku, and Kyusyu) and eleven categories for each five-year age group from 20 to 74.

### Ethical approval

This study was approved by the Institutional Review Board of the Institute of Economics Studies, Keio University (No. 21009R) and all participants provided informed consent.

### Patient and Public Involvement statement

There was no patient and public involvement in this study.

### Definitions of variables

#### (1) Vaccine-related questions

To measure vaccine hesitancy, we asked respondents their vaccination intentions, based on response options: already vaccinated, willing to vaccinate, undecided, and unwilling to vaccinate. Following the definition of vaccine hesitancy [2], we operationally defined those who were undecided and unwilling to vaccinate as vaccine hesitant.

In instances where respondents hesitated to vaccinate, we additionally asked them about the reasons for the unacceptance of a vaccine and asked them to indicate if each reason mattered to them. Referring to previous investigations [33, 34], we identified 18 items, such as concerns about the vaccine’s side effects, safety, efficacy, and other reasons.

#### (2) Independent variables

To account for the factors associated with vaccine hesitancy, as indicated by previous findings [3, 8-11], we obtained demographic, socioeconomic, health-related, and psychological information on each respondent.

The demographic and socioeconomic status of respondents included information on age, gender, co-resident family members, occupation, education, and income. Respondents living with members with chronic illnesses, aged 65 or over, and aged 11 or younger may be more likely to vaccinate because they are considered vulnerable to infection or not eligible for COVID-19 vaccination in Japan. In terms of occupation, we used three categories of essential healthcare workers, frontline essential workers, and other workers, following existing definitions [35]. Educational attainment of respondents included three categories of high school or lower, junior college or vocational school, and university or higher. Income refers to annual household income, obtained as the median value in 19 ranges.

We also used two health measures of self-rated health and depressive symptoms measured by the Kessler Psychological Distress Scale (K10), which have been validated by previous studies [36, 37]. Higher scores indicate better health for the former scale, whereas lower scores indicate worse health for the latter.

Finally, we used the following items to measure psychological and behavioural factors. Time and risk preferences, which relate to individuals’ responses to uncertain risks of vaccination and infection, were measured in the following two ways. The time preference was measured by the relative value of a larger later gain (13 months later) instead of a smaller immediate rewards (i.e. JPY 10,000 one month later) [38]. The risk preference was obtained as the sum of the responses to seven questions on risk attitudes measured using a seven-point Likert scale [39], which indicate higher scores representing more risk-loving. To measure respondents’ perceived seriousness of COVID-19, we used the Fear of COVID-19 Scale [40, 41]. We defined respondents’ numeracy as the number of correct answers to the same three questions proposed in a previous study [42].

### Empirical strategy

To assess the determinants of vaccine hesitancy, the reasons for hesitating vaccination, and efficacies of the relaxation of public health restrictions on vaccine acceptance, we conducted the following three analyses.

#### (1) Determinants of vaccine hesitancy

We evaluated the association between vaccine hesitancy and its determinants, including demographic, socioeconomic, health, and psychological factors. In the base model, we analysed the association using a logit model with a dichotomised outcome (i.e. unwilling to vaccinate or undecided vs. willing to vaccinate or already vaccinated). To test the robustness of the results, we assessed the association using a three-level nominal outcome (unwilling to vaccinate vs. undecided vs. willing to vaccinate or already vaccinated).

#### (2) Reasons for vaccine hesitancy

We investigated the determinants of the reasons for vaccine hesitancy by analysing the association between demographic, socioeconomic, health, and psychological factors and each reason given by the individuals hesitating to vaccinate. We estimated the marginal effects of the factors for each reason using a logit model.

#### (3) Conjoint analysis: vaccine passport

To evaluate the association between the relaxation of public health restrictions by vaccine passports and vaccine acceptance, we utilised a conjoint experimental design [18, 43]. The conjoint experiment is useful in assessing the effects of varied attributes at different levels, reducing the number of necessary assignments using an orthogonal table.

Using this design, in a hypothetical situation, we asked each respondent whether they would vaccinate, assuming that some or all four public health restrictions are relaxed. The four public health restrictions included travel across prefectures, dining out at night, joining gatherings and events, and going out without face masks, which correspond to government requests in Japan.

Without the design, we would need to assign 16 (= 2^4^) questions to assess each attribute of vaccine passports to each respondent; however, we reduced assignments by half, as shown in Table 1.

**Table 1.** Conjoint experimental design

|  | Available relaxations of restrictions by vaccine passports |  |  |  | Vaccine intensions |
| --- | --- | --- | --- | --- | --- |
|  | Travel across prefectures | Dining out after 8 p.m. | Joining gatherings and events | Going out without face masks | Yes / No |
| Pattern A | x | x | x | x |  |
| Pattern B | x | x | ○ | ○ |  |
| Pattern C | x | ○ | x | ○ |  |
| Pattern D | x | ○ | ○ | x |  |
| Pattern E | ○ | x | x | ○ |  |
| Pattern F | ○ | x | ○ | x |  |
| Pattern G | ○ | ○ | x | x |  |
| Pattern H | ○ | ○ | ○ | ○ |  |

In the conjoint experiment, all respondents provided their vaccination intentions for each hypothetical vaccine passport. To account for potential non-random variance across respondents arising from repeated measures, we fitted population-average panel-data models using the method of generalised estimating equations [44], estimating robust standard errors, and considering logit models with binominal distributions of outcomes.

All analyses were conducted using Stata MP, version 17.1 (StataCorp LLC, College Station, United States of America).

## Results

### Descriptive statistics

Table 2 summarises the descriptive statistics of the sample. Of a total of 5,000 respondents, about 30% hesitated to vaccinate (i.e. unwilling to vaccinate: 12.5% and undecided: 17.9%). Vaccination intentions and status can change over time due to various factors, such as infection situation and vaccine availability. At the time of the survey, about 38% of the Japanese population had at least one dose of the COVID-19 vaccine[5], health professionals and older adults being prioritized. The proportion of the vaccinated population was almost identical to that of our sample (36.6%), indicating that our sample reflects the Japanese context well.

**Table 2.** Descriptive statistics (n = 5,000)

| Variable | Mean or proportion | Standard deviation |
| --- | --- | --- |
| Vaccine intentions: No | 12.5% |  |
| Undecided | 17.9% |  |
| Yes | 33.1% |  |
| Already vaccinated | 36.6% |  |
| Age | 48.40 | 14.80 |
| Female | 50.3% |  |
| Co-residence: Aged 65 or older | 33.3% |  |
| Aged 11 or younger | 14.4% |  |
| Chronic illness | 25.0% |  |
| Occupation: Healthcare worker | 6.4% |  |
| Frontline essential workers | 26.3% |  |
| Other occupations | 31.8% |  |
| Not employed (Ref.) | 35.5% |  |
| Education: High school or lower (Ref.) | 31.2% |  |
| Junior college or vocational | 19.5% |  |
| University or higher | 49.2% |  |
| Household income (million JPY) | 5.60 | 3.78 |
| Self-rated health | 3.52 | 1.02 |
| K10 depression scale | 17.95 | 9.18 |
| Numeracy | 1.58 | 0.78 |
| Time preference | 21.08 | 16.21 |
| Risk preference | 27.85 | 8.3 |
| Fear of COVID-19 | 19.63 | 5.46 |
| Residential area: Hokkaido | 4.6% |  |
| Tohoku | 5.9% |  |
| Kanto | 26.1% |  |
| Chubu | 14.9% |  |
| Kinki | 14.3% |  |
| Chugoku | 19.3% |  |
| Shikoku | 6.7% |  |
| Kyushu | 8.0% |  |

Table 3 presents the reasons for vaccine hesitancy among the individuals who hesitate to vaccinate. The most major concerns were the vaccine’s side effects and safety (87%), as well as other reasons related to vaccine safety, preference, and mistrust being commonly reported by respondents.

**Table 3.** Reasons for vaccine hesitancy

| Reasons | % |
| --- | --- |
| Concern about side effects and safety of the vaccine | 87% |
| Plan to wait and see if it is safe and may get it later | 79% |
| Concern that the vaccine is being developed too quickly | 73% |
| Plan to use masks/other precautions instead | 69% |
| Do not trust the government | 67% |
| Do not like vaccines | 63% |
| Do not like needles | 48% |
| Do not know I needed a vaccine against COVID-19 | 45% |
| The vaccine could give me COVID-19 | 37% |
| The vaccine will not work | 31% |
| I will not need vaccinate because vaccination of other people will establish herd immunity | 29% |
| Vaccination site is far | 28% |
| COVID-19 is not a serious illness | 26% |
| Too busy to visit a vaccination site | 25% |
| Had COVID-19 and should be immune | 11% |
| Doctor has not recommended a COVID-19 vaccine to me | 11% |
| Pregnant | 7% |
| For religious reasons | 5% |

Note: Percentages among 1,518 respondents hesitating vaccination.

### Determinants of vaccine hesitancy

Table 4 shows the determinants of vaccine hesitancy. Compared to those aged 45– 49, younger people aged 25–44 were likely to hesitate to vaccinate, resulting in estimated odds ratios (ORs) ranging between 1.32 and 1.87, with 95% confidence intervals (CI) ranging from 1.01 to 2.44. Meanwhile, older adults aged 55–74 tended to accept vaccination, showing estimated ORs between 0.17 and 0.67 with 95% CI from 0.11 to 0.89. Female respondents tended to express vaccine hesitancy more than their male counterparts (OR: 1.18, 95% CI: 1.02 – 1.37). Additionally, those living with older adults and members with chronic illness tended to accept vaccination with higher probabilities of 16–44% compared to their counterparts not living with these population categories.

**Table 4.**
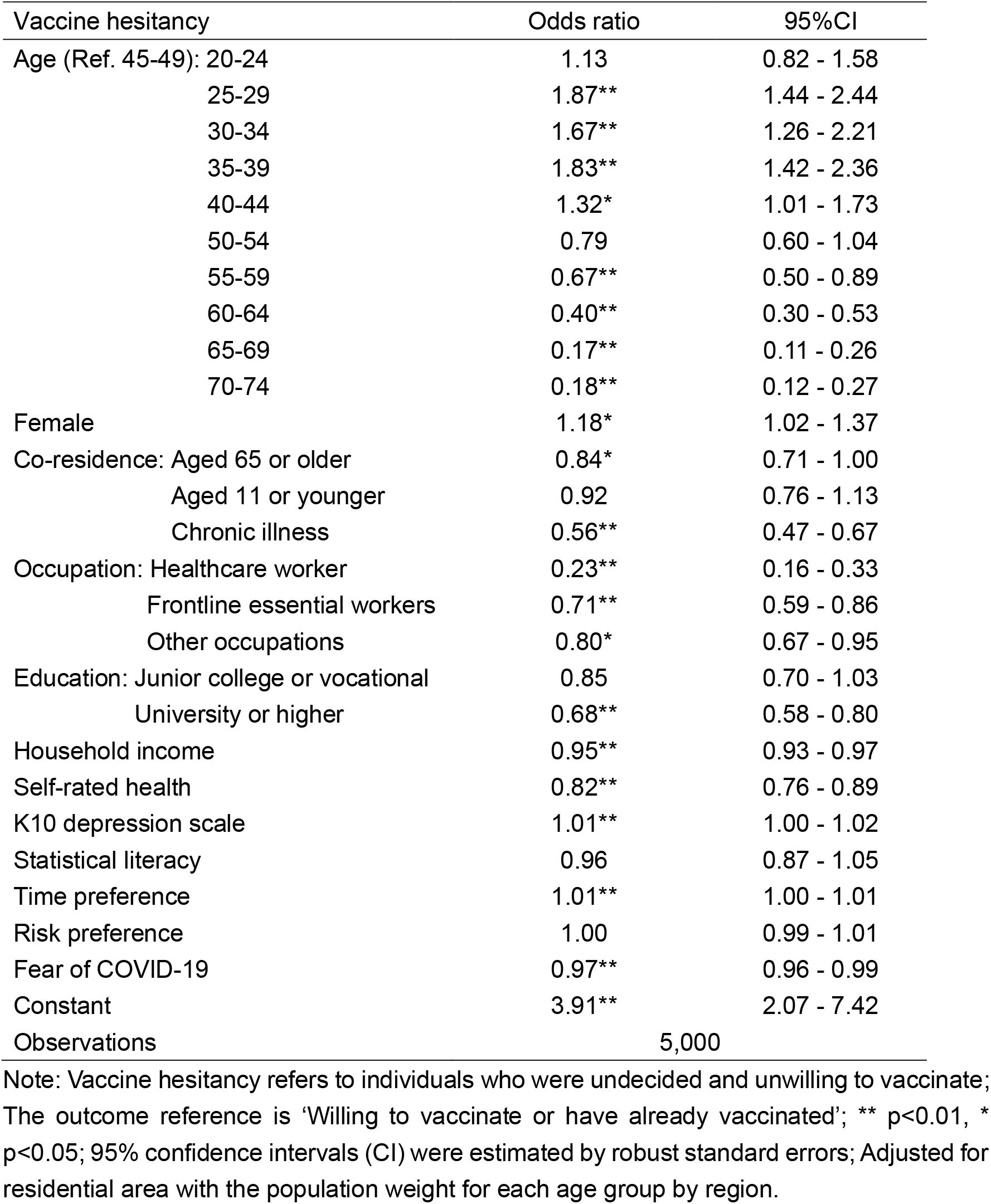
Determinants of vaccine hesitancy

| Vaccine hesitancy | Odds ratio | 95%CI |
| --- | --- | --- |
| Age (Ref. 45-49): 20-24 | 1.13 | 0.82 - 1.58 |
| 25-29 | 1.87** | 1.44 - 2.44 |
| 30-34 | 1.67** | 1.26 - 2.21 |
| 35-39 | 1.83** | 1.42 - 2.36 |
| 40-44 | 1.32* | 1.01 - 1.73 |
| 50-54 | 0.79 | 0.60 - 1.04 |
| 55-59 | 0.67** | 0.50 - 0.89 |
| 60-64 | 0.40** | 0.30 - 0.53 |
| 65-69 | 0.17** | 0.11 - 0.26 |
| 70-74 | 0.18** | 0.12 - 0.27 |
| Female | 1.18* | 1.02 - 1.37 |
| Co-residence: Aged 65 or older | 0.84* | 0.71 - 1.00 |
| Aged 11 or younger | 0.92 | 0.76 - 1.13 |
| Chronic illness | 0.56** | 0.47 - 0.67 |
| Occupation: Healthcare worker | 0.23** | 0.16 - 0.33 |
| Frontline essential workers | 0.71** | 0.59 - 0.86 |
| Other occupations | 0.80* | 0.67 - 0.95 |
| Education: Junior college or vocational | 0.85 | 0.70 - 1.03 |
| University or higher | 0.68** | 0.58 - 0.80 |
| Household income | 0.95** | 0.93 - 0.97 |
| Self-rated health | 0.82** | 0.76 - 0.89 |
| K10 depression scale | 1.01** | 1.00 - 1.02 |
| Statistical literacy | 0.96 | 0.87 - 1.05 |
| Time preference | 1.01** | 1.00 - 1.01 |
| Risk preference | 1.00 | 0.99 - 1.01 |
| Fear of COVID-19 | 0.97** | 0.96 - 0.99 |
| Constant | 3.91** | 2.07 - 7.42 |
| Observations | 5,000 |  |

Note: Vaccine hesitancy refers to individuals who were undecided and unwilling to vaccinate; The outcome reference is 'Willing to vaccinate or have already vaccinated'; \*\* p<0.01, \* p<0.05; 95% confidence intervals (CI) were estimated by robust standard errors; Adjusted for residential area with the population weight for each age group by region.

Socioeconomic factors are also associated with vaccine hesitancy. Healthcare workers, frontline essential workers, and those performing paid work were likely to be non-vaccine-hesitant compared to non-employed individuals: the former two groups were more likely to accept vaccination, showing ORs of 0.23 (95% CI: 0.16–0.33) and 0.71 (95% CI: 0.59–0.86), respectively. Furthermore, higher education and income were associated with a lower likelihood of being vaccine-hesitant.

Those with poorer health, measured by self-rated health and the K10 depression scale, were less likely to hesitate to vaccinate by 18% (95% CI: 0.76–0.89) and 1% (95% CI: 1.00–1.02), respectively. Psychological and behavioural factors such as time preference and fear of COVID-19 were also predictors of vaccine hesitancy.

To check the robustness of the results, we decomposed vaccine hesitancy into two groups: those unwilling to vaccinate and those who have not yet decided. Similar results were observed for the findings estimated from the binary outcomes (Appendix Table A-1).

### Reasons for vaccine hesitancy

In Figure 1 and 2, we present the results of the analyses on the reasons related to the side effects and safety of the vaccine and vaccine mistrust, which were the most common. While we did not find remarkable heterogeneity across most factors, a higher numeracy was associated with vaccine hesitancy due to concerns about vaccine side effects and safety.

**Figure 1.**
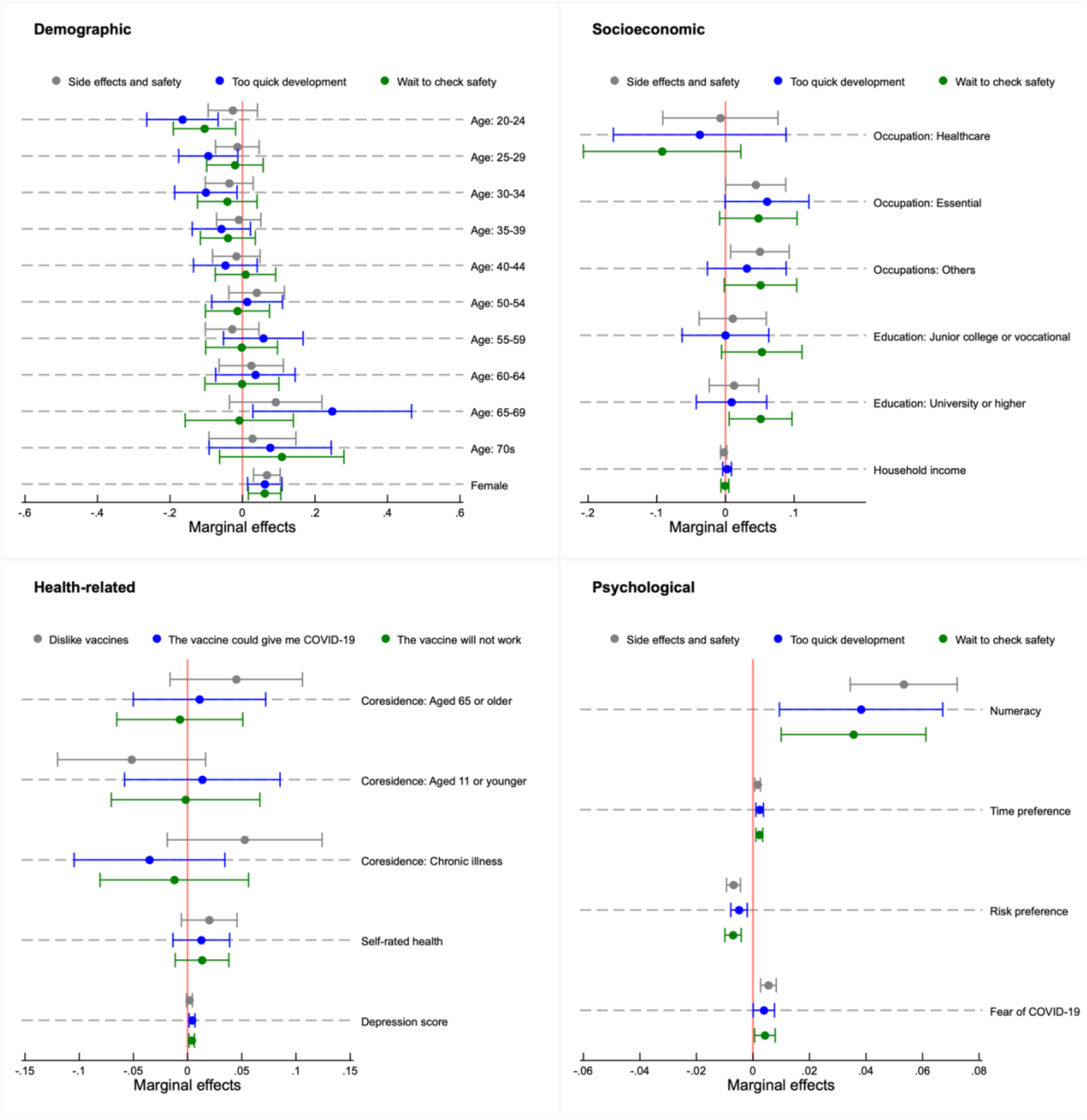
Determinants of reasons for vaccine hesitancy: Side effects and safety. Note: Analyses among individuals who were undecided and unwilling to vaccinate (n= 1,518); Markers represent marginal effects with error bars showing 95% confidence intervals estimated by robust standard errors; Adjusted for residential area with the population weight for each age group.

**Figure2.**
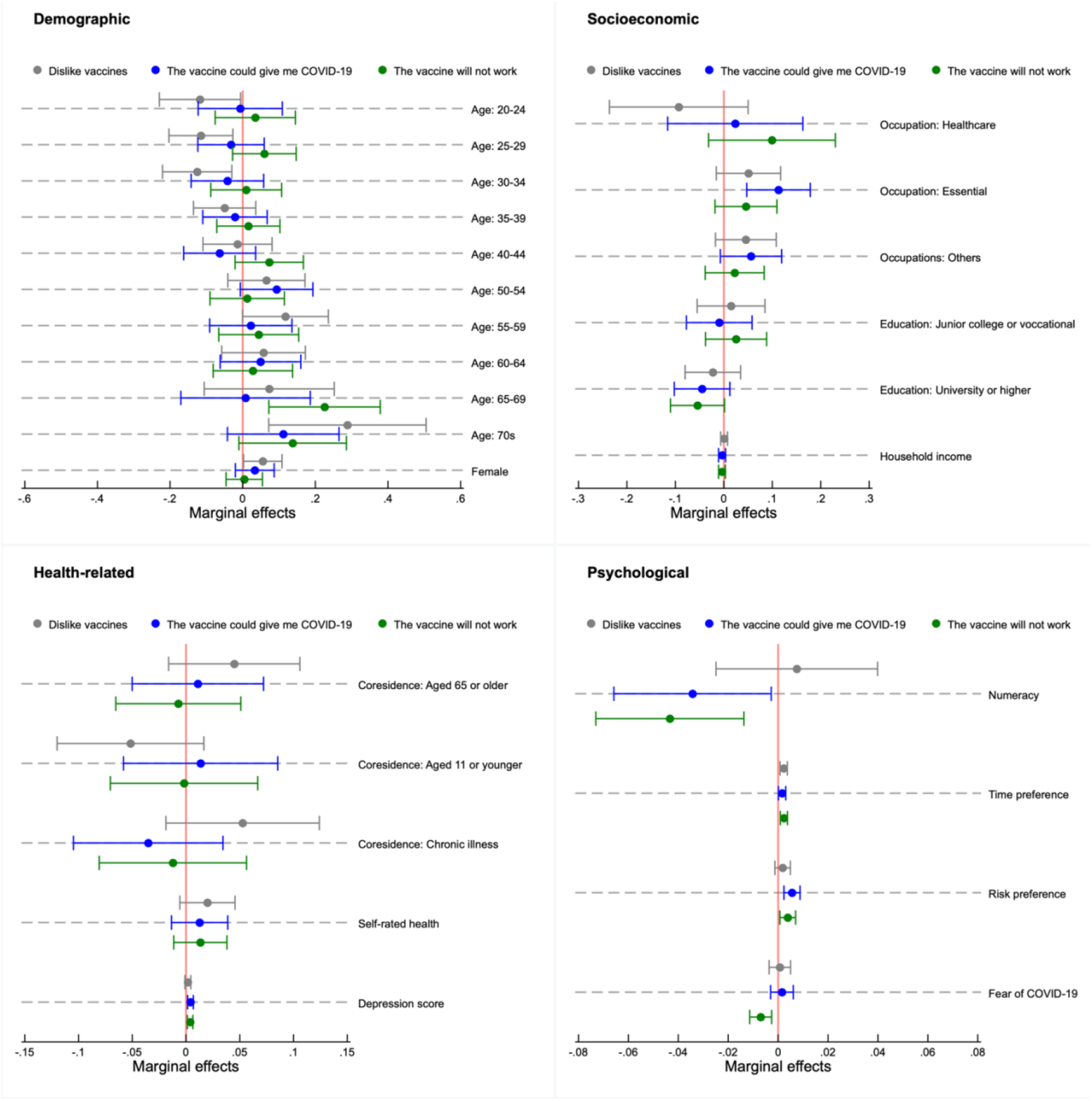
Determinants of reasons for vaccine hesitancy: Vaccine mistrust. Note: Analyses among individuals who were undecided and unwilling to vaccinate (n= 1,518); Markers represent marginal effects with error bars showing 95% confidence intervals estimated by robust standard errors; Adjusted for residential area with the population weight for each age group.

Additionally, the results for other reasons are shown in Appendix Figures A-1, A-2, A-3, and A-4. Again, we did not find systematic trends in the determinants of the reasons for vaccine hesitancy.

### Effectiveness of vaccine passport

From the conjoint experiment, we observed that 45% of all the vaccine hesitant intended to accept vaccination when all public health restrictions were relaxed, while 18% intended so if no restrictions were relaxed.

In Figure 3, we present the estimation results for the association between the relaxation of each public health restrictions and vaccine acceptance, suggesting that relaxing all restrictions were effective in increasing vaccine acceptance by 4–10%. In particular, the relaxation of travel restriction across prefectures was the most effective, showing a 10% increase (95% CI: 9-11%) in vaccine acceptance, if permitted.

**Figure 3.**
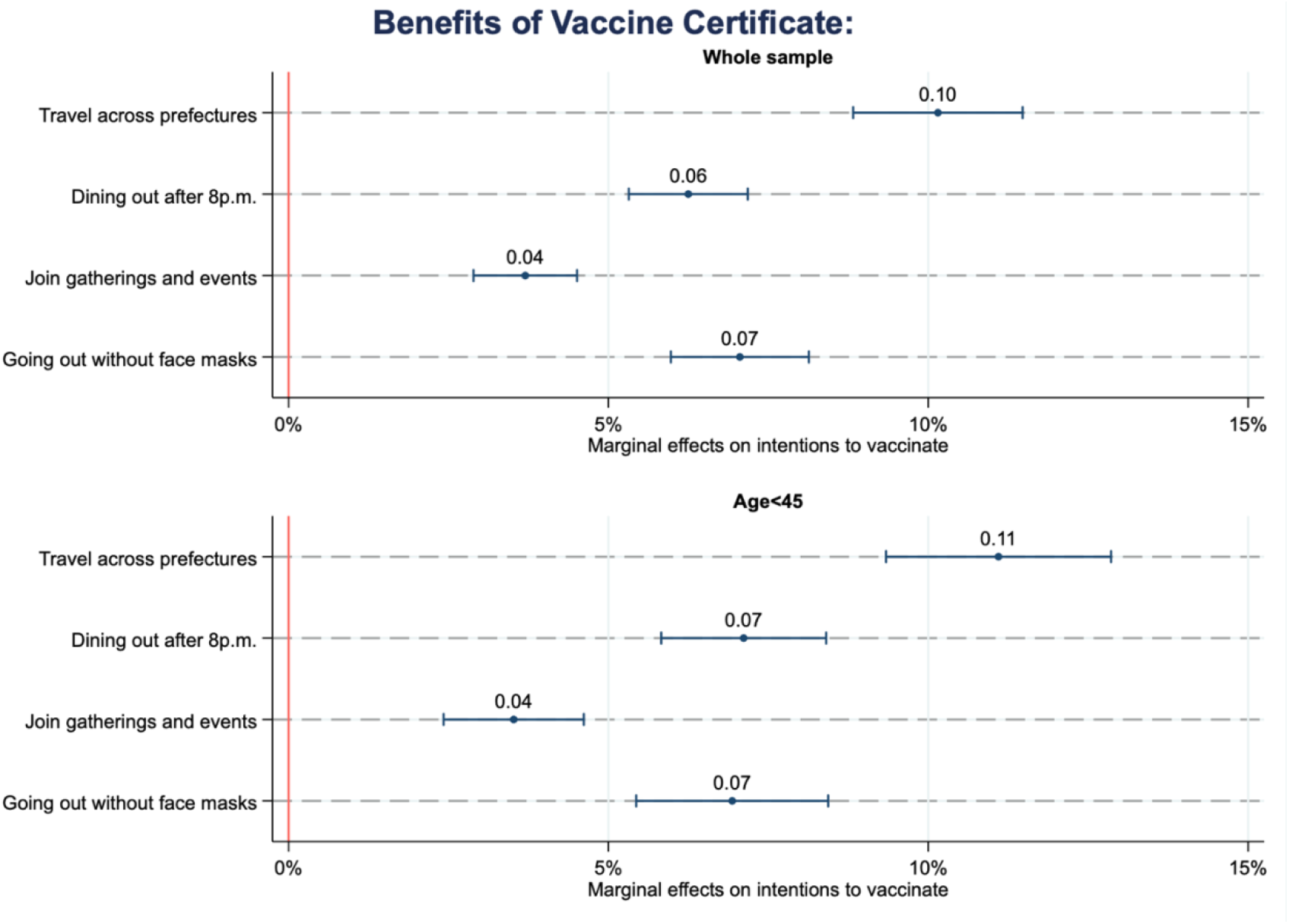
Effectiveness of vaccine passport. Note: Estimates among individuals who were undecided and unwilling to vaccinate (n= 1,518 for all age groups and n= 884 for those aged 45 or younger); Adjusted for age, gender, co-resident family members, occupation, education, income, health status, statistical literacy, time preference, risk preference, fear of COVID-19, residential area, and vaccine attributes with the population weight for each age group by region; Values and markers are marginal effects with bars representing 95% confidence intervals estimated by robust standard errors; Full results are available upon request.

Moreover, we analysed the potential heterogeneity among younger people aged 44 or younger, who were more likely to be vaccine-hesitant in our previous analysis. We found that the results remained unchanged and these policies were effective for younger people.

Additionally, we conducted the following robustness tests. First, we excluded the respondents whose choices may have been nontransitive. Some individuals preferred not to vaccinate when an additional relaxation was offered, although they expressed their willingness to vaccinate with fewer options. Although this may suggest that they did not prefer to ease certain restrictions regardless of vaccination status, we re-analysed the association by excluding them. However, the results remained unchanged (Appendix Figure A-5). Second, we separately analysed respondents who were undecided and unwilling to vaccinate. Although marginal effects among those unwilling to vaccinate become smaller while the estimates undecided respondents became larger, we found that the relaxation of public health restrictions were evidently effective to increase vaccine acceptance (Appendix A-6). Third, we included respondents who intended to vaccinate earlier or have already been vaccinated, and the same results were still observed (Appendix Figure A-7). Next, from among the individuals hesitating to vaccinate, we excluded those who provided a uniform answer to all options (i.e. all yes or no), the results remaining unchanged (Appendix Figure A-8). Finally, to relax the assumption of the independence of irrelevant alternatives, we estimated our main model by a multilevel mixed-effects logistic regression and confirmed the results were still robust (Appendix Figure A-9).

### Stated and revealed preferences

About four months after this survey (i.e. between 10th and 20th of November 2021), we conducted a follow-up survey and obtained 4,367 responses out of 5,000 participants in the first wave (87.3%). For respondents who had not vaccinated yet at the first survey, we present descriptive statistics on vaccination status at the follow-up survey (Table 5). More than 90% of respondents who intended to vaccinate actually receive it while smaller proportions among those undecided and unwilling to vaccinate did so. About 29% of undecided and 69% of unwilling individuals remain vaccine hesitant in the follow-up survey.

**Table 5.**
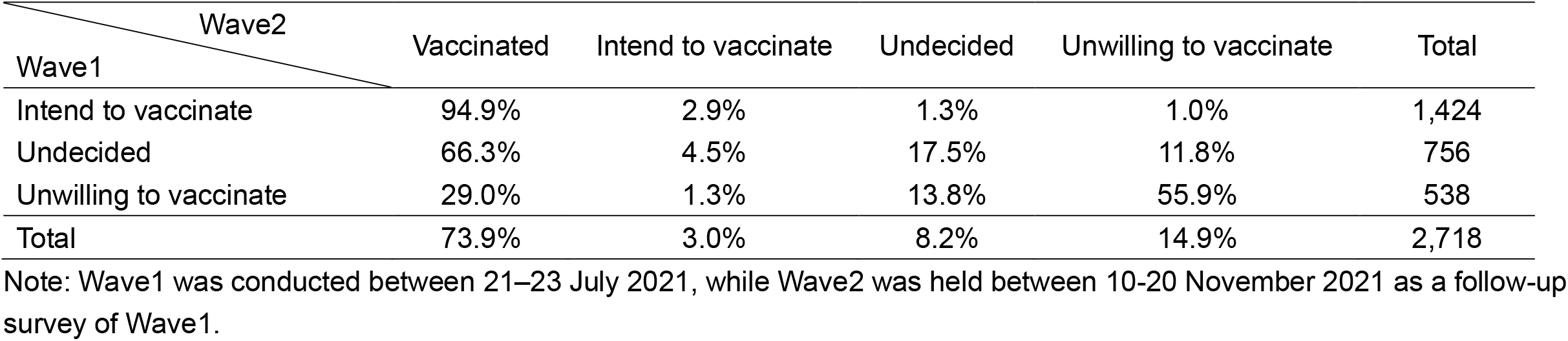
Stated and revealed preferences

## Discussion

This study primarily assessed whether easing public health restrictions after vaccination increases vaccine acceptance, as well as investigated determinants of vaccine hesitancy and reasons for vaccine hesitancy. As the first study to explore the effectiveness of the relaxation of public health restrictions, by decomposing what can be permitted by vaccination, we obtained three main findings. First, in line with previous findings [3, 8-11], our analysis suggests that demographic, socioeconomic, health-related, and psychological factors predict vaccine hesitancy. In particular, younger age seems to be the strongest predictor of vaccine hesitancy, while other factors, such as gender and socioeconomic factors, were also associated with vaccine hesitancy. Second, concerns about the side effects and safety of the COVID-19 vaccine, as well as mistrust of vaccines and the government in general, were dominant reasons for vaccine hesitancy. Meanwhile, we did not observe remarkable heterogeneity in the association between age, which was found to be a strong predictor of vaccine hesitancy, and the reasons for vaccine hesitancy. Third, we found that vaccination acceptance increases by easing public health restrictions, especially travel restrictions across prefectures. This result was particularly evident among younger people, who had higher probabilities of vaccine hesitancy than their older counterparts.

One reason why younger people tend to hesitate to vaccinate is the expected balance between the costs and benefits of vaccination, as predicted by the health belief model and economic theory [7, 13]. Considering that younger people are less likely to develop severe COVID-19 symptoms than older people [16] and given the higher likelihood of the side effects of the vaccine (e.g. headache and fatigue) among them [17], they could decide not to vaccinate from a utilitarian perspective. While we did not observe remarkable trends for the association between age and the reasons for hesitating to vaccinate, vaccine safety and side effects were the most common reasons, which has also been reported by other studies [33, 34]. Moreover, we found that statistical numeracy predicts vaccine hesitancy due to the concerns about vaccine safety and side effects. This may suggest that, being related to prospect theory, statistical capacity is related to inconsistent preferences and overestimating health losses of vaccination [45].

Our study also suggests that vaccine passports, which allow citizens freedom in their daily lives, could increase vaccine acceptance. In many countries, including Japan, individuals are subject to containment and closure policies by governments to curb the spread of the virus, which requires them to avoid non-essential activities, such as eating out, travelling, and mass gatherings. As stay-at-home orders and social distancing behaviours can deteriorate citizens’ well-being and mental health through distress, boredom, loneliness, and social isolation [46], eliminating public health restrictions and returning to a normal life may be what many citizens are eager to attain.

Based on our findings, several policy implications can be drawn. First, information campaigns to convey accurate messages are extremely important to enhance the understanding and remove vaccine mistrust. Utilising behavioural insights, better designs on how to best communicate with people to enhance vaccine uptake are considered [23, 24]. Second, emphasising benefits other than health (e.g. the relaxation of public health restrictions), if applicable, may enhance vaccine acceptance. Even under the concerns about breakthrough infections, the overall public health benefits may be in surplus if and only if a rise in vaccine acceptance largely reduces severe symptoms and the mortality rate from infection given the confirmed safety of the vaccine by contributing to the establishment of herd immunity against COVID-19. The continuous evaluations and careful consideration of the efficacy, duration of effectiveness, and side effects of the vaccine, as well as potential public health impacts and ethical issues of vaccine passports are indispensable. With the uncertain duration of vaccine efficacy and the efficacy against new virus variants, it would be realistic to issue vaccine passports for a limited time, maintaining moderate infectious control measures. Furthermore, these types of passports must not be used to discriminate and eliminate those not vaccinating from society, allowing them to use alternative services, such as a certificate for a negative COVID-19 test result.

In this study, we first provided evidence on the effectiveness of vaccine passports to relax the public health restrictions, decomposing the activities allowed by passports, on reducing vaccine hesitancy. Nevertheless, several limitations should be noted due to caveats. First, our study was based on a hypothetical experiment and not a real situation. Therefore, actual behaviour may diverge from the survey responses. Notwithstanding this limitation, our findings should still be helpful, based on previous reports that more than 80% of the stated and revealed preferences corresponded [47, 48]. Second, our findings are based on a single survey, in which the sample was obtained from registered panels that may not be identical to the general public. Although we utilised weights estimated by population structures by region, other factors than these could not be representative. Also, vaccination intentions may fluctuate due to the pandemic situation and the timing of the survey. Furthermore, our results may not be applicable in other countries, since the pandemic situation, government responses to the pandemic, and reasons for vaccine hesitancy can vary across countries. In Japan, compared to Western countries, the COVID-19 mortality rate is much lower [49], and government responses to the pandemic are less stringent [50]; hence, the attitude of the Japanese population toward the vaccine and vaccine passports availability may differ from other countries. Therefore, intertemporal and cross-national evidence needs to be accumulated through further studies.

## Conclusions

In conclusion, this work offers encouraging findings regarding the vaccination intentions of the Japanese people. Although some individuals hesitate to get vaccinated against COVID-19, this can be reduced by mitigating concerns about vaccine safety and side effects throughout appropriate and effective information campaigns. Additionally, the relaxation of public health restrictions, such as travel across prefectures, wearing face masks, and dining out at night, is effective in enhancing vaccine acceptance. To assist the progress toward herd immunity, the feasibility of vaccine passports needs to be sufficiently assessed by taking the ethical issues of the passports and public health impacts of the relaxation of restrictions into careful consideration.

## Data Availability

The data used in this study are not available as we did not obtain the agreement of the participants to make the data open.

## Acknowledgements

We thank Dr Rei Goto, Dr Hirotaka Kato, Mr Shingo Kasahara, Mr Tatsunari Miyayama, and Ms Tomomi Maeda at Keio University for their helpful feedback. We are also grateful to Ms Haruka Umijima at Keio University for her administrative assistance to conduct the study.

## a. Contributorship statement

Okamoto, Kamimura, and Komamura conceptualised the study and were engaged in the data collection. Okamoto and Kamimura conducted analyses, which was further refined and finalised by Okamoto. Komamura also provided critical comments to refine analysis. Okamoto prepared a first draft, which was reviewed by Kamimura and Komamura.

## d. Data sharing statement

The data underlying this article cannot be shared publicly for the privacy of individuals that participated in the study. We did not obtain consents from respondents to make the data open.

## Supplementary Material for

### Appendix Tables

**Appendix Table A-1.** Determinants of vaccine hesitancy

| Vaccine hesitancy | Will not vaccinate |  | Undecided |  |
| --- | --- | --- | --- | --- |
|  | Relative risk ratios | 95%CI | Relative risk ratios | 95%CI |
| Age (Ref. 45-49): 20-24 | 1.48 | 0.95 - 2.30 | 0.94 | 0.63 - 1.41 |
| 25-29 | 1.97** | 1.36 - 2.85 | 1.83** | 1.35 - 2.47 |
| 30-34 | 2.08** | 1.40 - 3.09 | 1.47* | 1.07 - 2.03 |
| 35-39 | 2.22** | 1.55 - 3.18 | 1.63** | 1.22 - 2.19 |
| 40-44 | 1.46 | 0.99 - 2.15 | 1.25 | 0.92 - 1.70 |
| 50-54 | 0.87 | 0.58 - 1.29 | 0.75 | 0.55 - 1.04 |
| 55-59 | 0.89 | 0.59 - 1.35 | 0.55** | 0.38 - 0.78 |
| 60-64 | 0.53** | 0.35 - 0.79 | 0.33** | 0.23 - 0.47 |
| 65-69 | 0.24** | 0.13 - 0.42 | 0.14** | 0.08 - 0.23 |
| 70-74 | 0.30** | 0.18 - 0.51 | 0.11** | 0.07 - 0.20 |
| Female | 1.08 | 0.88 - 1.32 | 1.26** | 1.06 - 1.51 |
| Co-residence: Aged 65 or older | 0.75* | 0.60 - 0.95 | 0.91 | 0.74 - 1.11 |
| Aged 11 or younger | 0.68* | 0.50 - 0.92 | 1.10 | 0.88 - 1.38 |
| Chronic illness | 0.53** | 0.41 - 0.68 | 0.59** | 0.47 - 0.73 |
| Occupation: Healthcare worker | 0.26** | 0.16 - 0.43 | 0.20** | 0.13 - 0.33 |
| Other essential workers | 0.56** | 0.43 - 0.73 | 0.83 | 0.67 - 1.04 |
| Other occupations | 0.70** | 0.55 - 0.89 | 0.88 | 0.71 - 1.09 |
| Education: Junior college or vocational | 0.82 | 0.63 - 1.07 | 0.87 | 0.69 - 1.10 |
| University or higher | 0.60** | 0.48 - 0.75 | 0.74** | 0.61 - 0.90 |
| Household income | 0.96* | 0.94 - 0.99 | 0.94** | 0.92 - 0.97 |
| Self-rated health | 0.79** | 0.71 - 0.88 | 0.85** | 0.78 - 0.92 |
| K10 depression scale | 1.01** | 1.00 - 1.03 | 1.01 | 1.00 - 1.02 |
| Statistical literacy | 0.85** | 0.75 - 0.96 | 1.05 | 0.94 - 1.17 |
| Time preference | 1.01* | 1.00 - 1.01 | 1.01* | 1.00 - 1.01 |
| Risk preference | 1.00 | 0.99 - 1.02 | 0.99 | 0.98 - 1.00 |
| Fear of COVID-19 | 0.95** | 0.93 - 0.97 | 0.99 | 0.98 - 1.01 |
| Constant | 3.01* | 1.25 - 7.22 | 1.35 | 0.64 - 2.85 |
| Observations | 5,000 |  |  |  |

Note: The outcome reference is 'Willing to vaccinate or have already vaccinated'; \*\* p<0.01, \* p<0.05; 95% confidence intervals (CI) were estimated by robust standard errors; Adjusted for residential area with the population weight for each age group by region.

### Appendix Figures

**Appendix Figure A-1.**
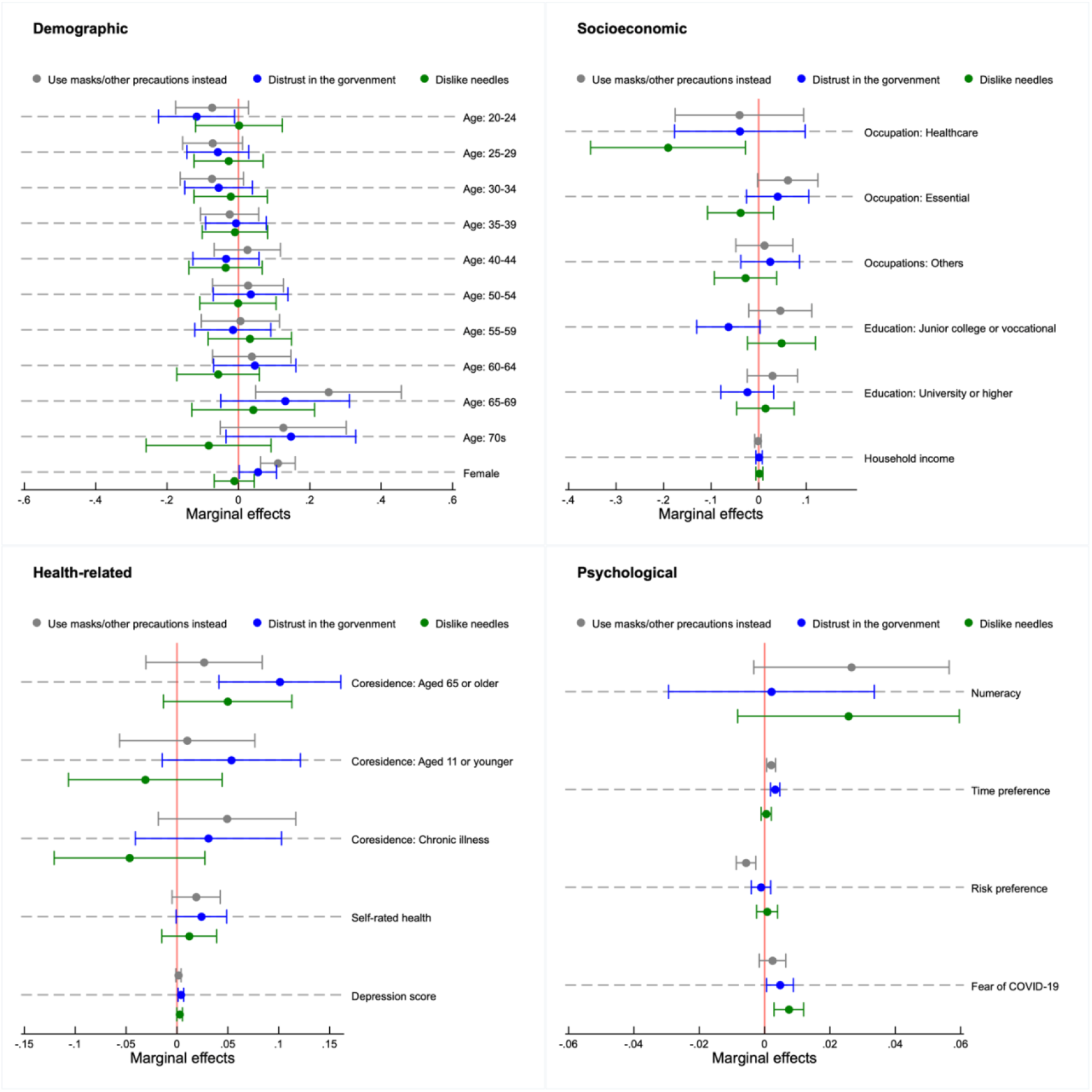
Determinants of reasons for vaccine hesitancy: Other mistrust. Note: Analyses among individuals who were undecided and unwilling to vaccinate (n= 1,518); Markers represent marginal effects with error bars showing 95% confidence intervals estimated by robust standard errors; Adjusted for residential area with the population weight for each age group.

**Appendix Figure A-2.**
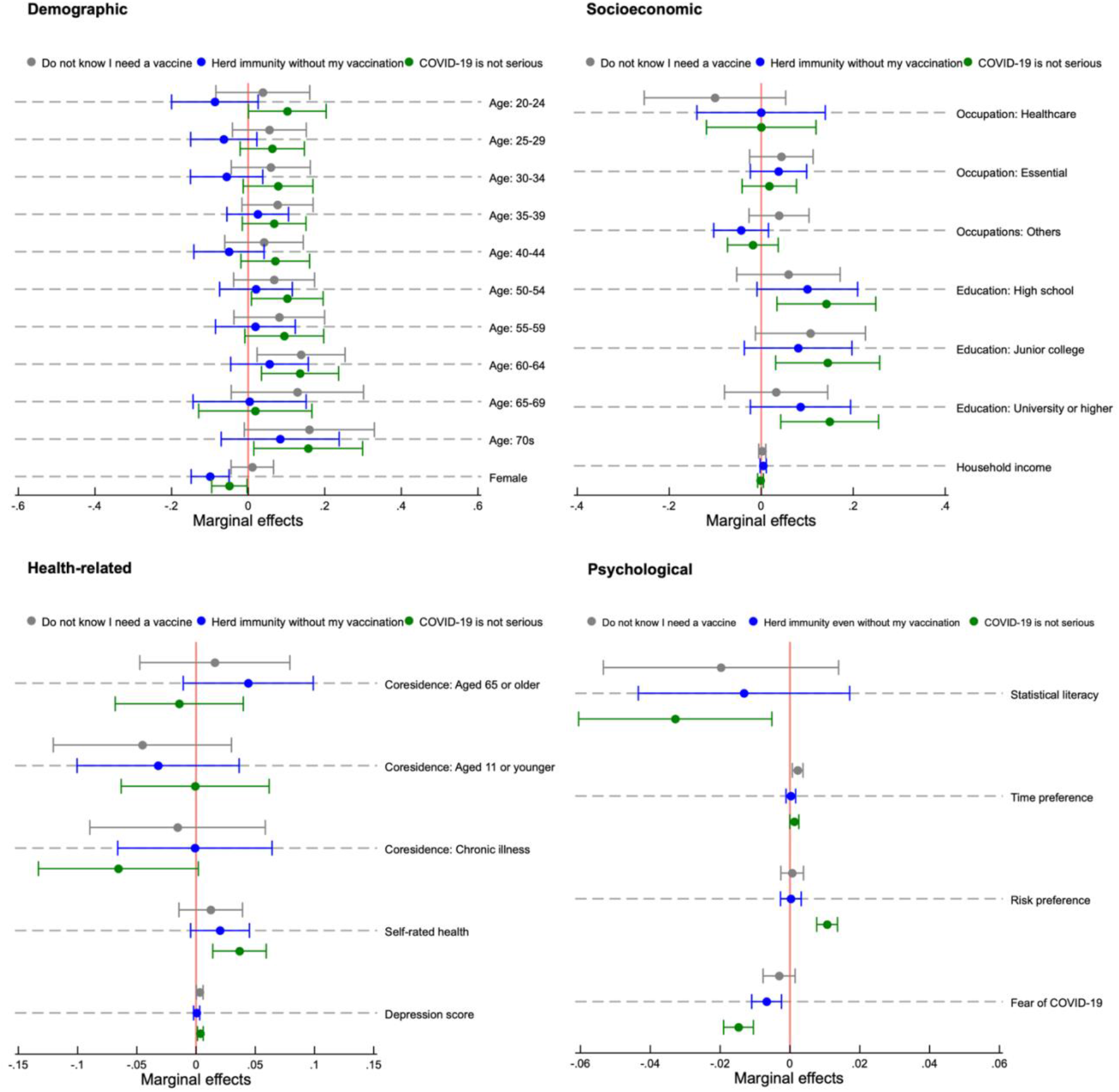
Determinants of reasons for vaccine hesitancy: Attitudes toward COVID-19. Note: Analyses among individuals who were undecided and unwilling to vaccinate (n= 1,518); Markers represent marginal effects with error bars showing 95% confidence intervals estimated by robust standard errors; Adjusted for residential area with the population weight for each age group;

**Appendix Figure A-3.**
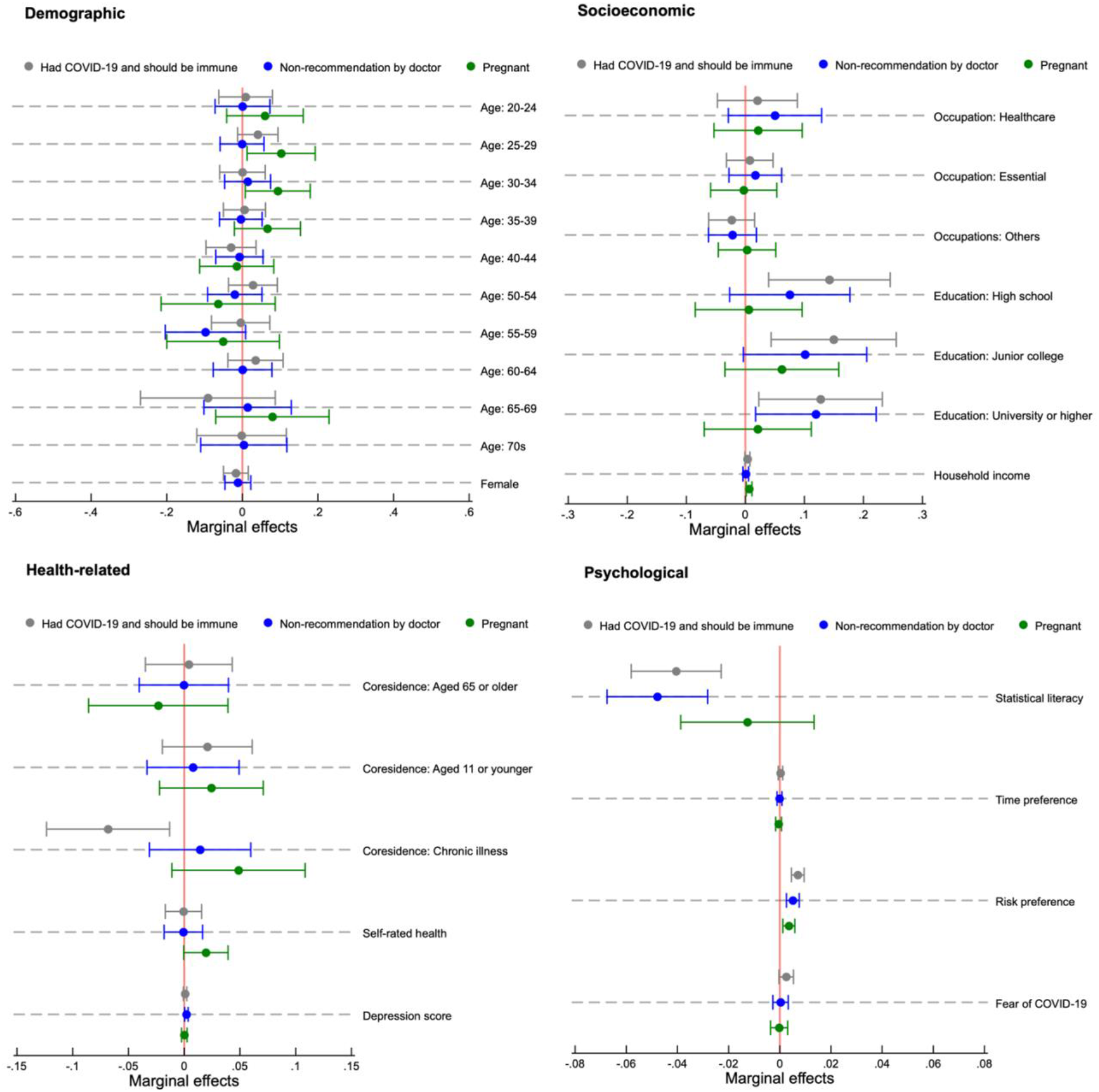
Determinants of reasons for vaccine hesitancy: Health-related reasons. Note: Analyses among individuals who were undecided and unwilling to vaccinate (n= 1,518); Markers represent marginal effects with error bars showing 95% confidence intervals estimated by robust standard errors; Adjusted for residential area with the population weight for each age group;

**Appendix Figure A-4.**
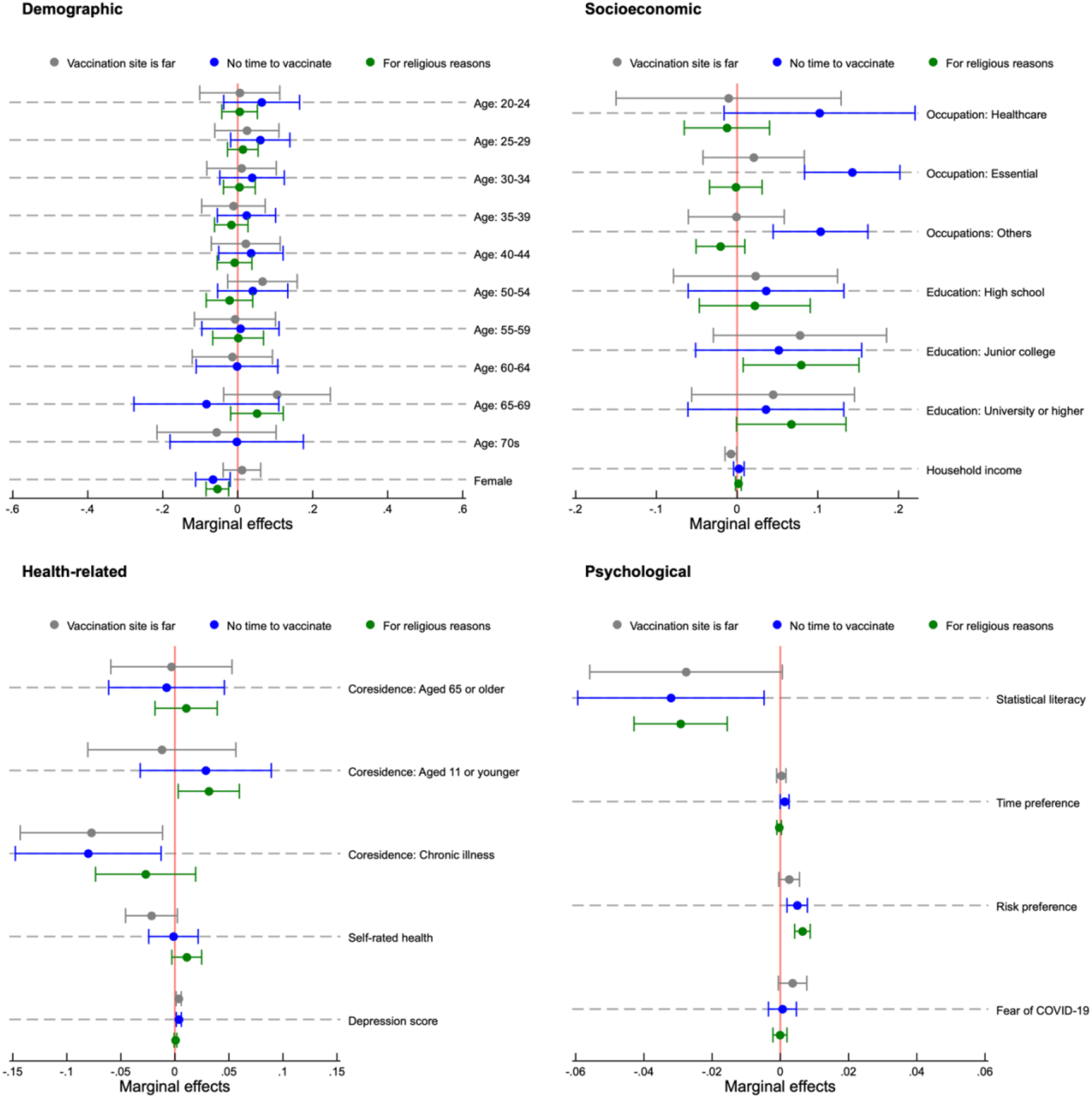
Determinants of reasons for vaccine hesitancy: Other reasons. Note: Analyses among individuals who were undecided and unwilling to vaccinate (n= 1,518); Markers represent marginal effects with error bars showing 95% confidence intervals estimated by robust standard errors; Adjusted for residential area with the population weight for each age group;

**Appendix Figure A-5.**
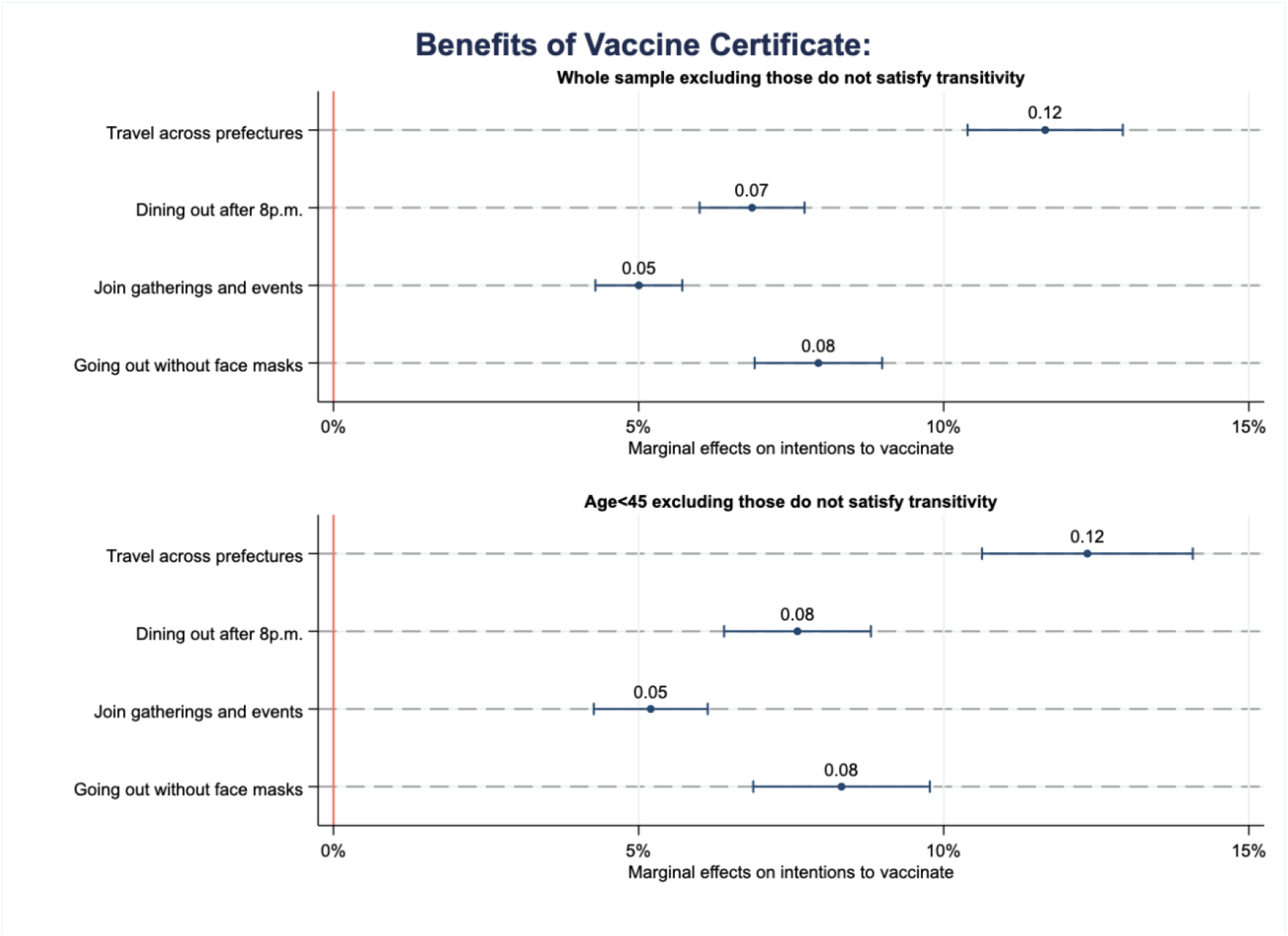
Vaccine passport: Exclude those with non-transitive preferences. Note: Estimates among individuals who were undecided and unwilling to vaccinate, excluding those who did not satisfy transitivity of the vaccine preference (n= 1,416 for all age groups and n= 815 for those aged 45 or younger); Adjusted for age, gender, co-resident family members, occupation, education, income, health status, statistical literacy, time preference, risk preference, fear of COVID-19, residential area, and vaccine attributes with the population weight for each age group by region; Values and markers are marginal effects with bars representing 95% confidence intervals estimated by robust standard errors; Full results are available upon request.

**Appendix Figure A-6.**
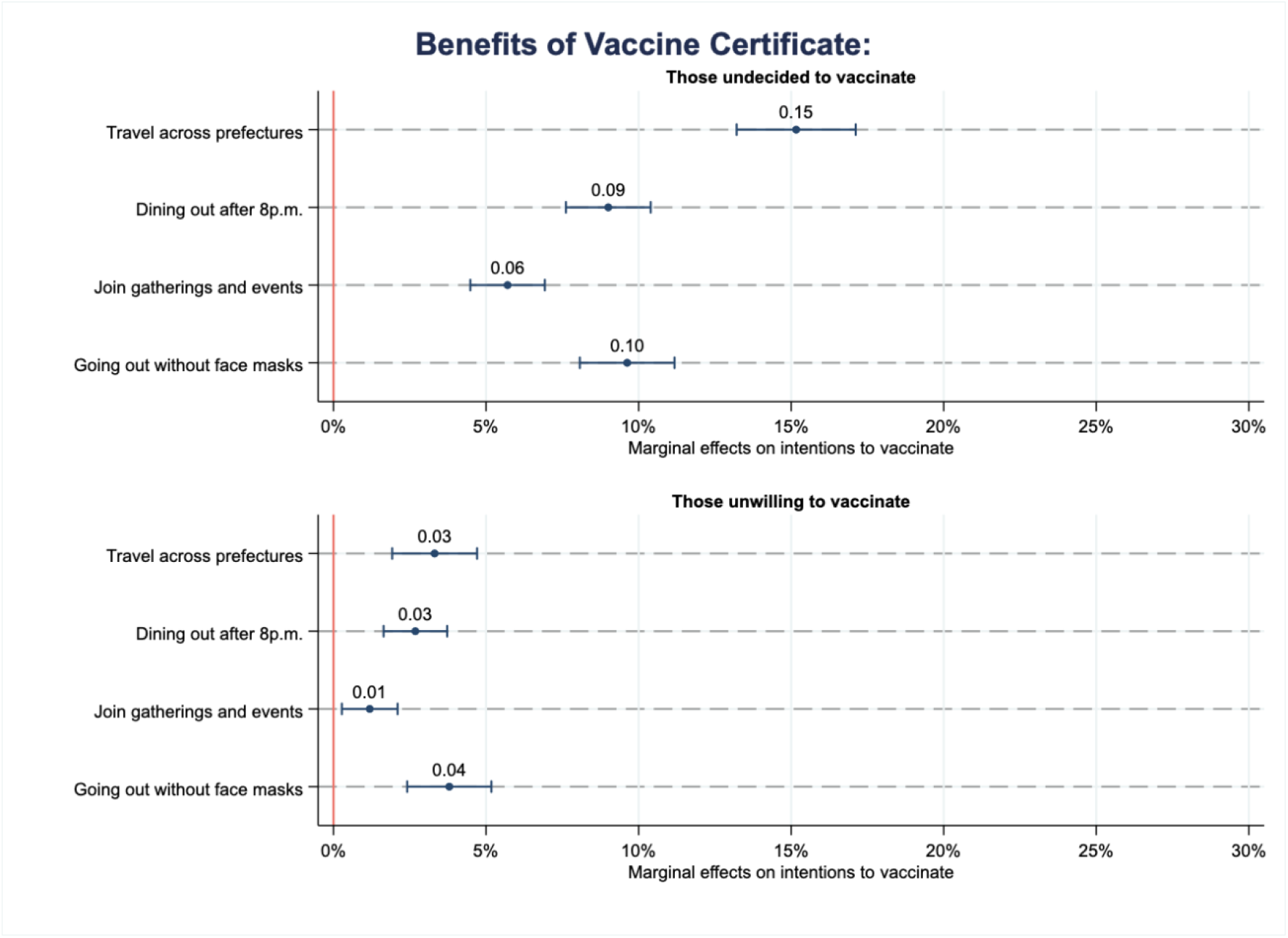
Vaccine passport: Separate analyses of those undecided and unwilling to vaccinate. Note: Estimates among individuals who were unwilling to vaccinate (n= 894 for those undecided to vaccinate and n= 624 for those unwilling to vaccinate); Adjusted for age, gender, co-resident family members, occupation, education, income, health status, statistical literacy, time preference, risk preference, fear of COVID-19, residential area, and vaccine attributes with the population weight for each age group by region; Values and markers are marginal effects with bars representing 95% confidence intervals estimated by robust standard errors; Full results are available upon request.

**Appendix Figure A-7.**
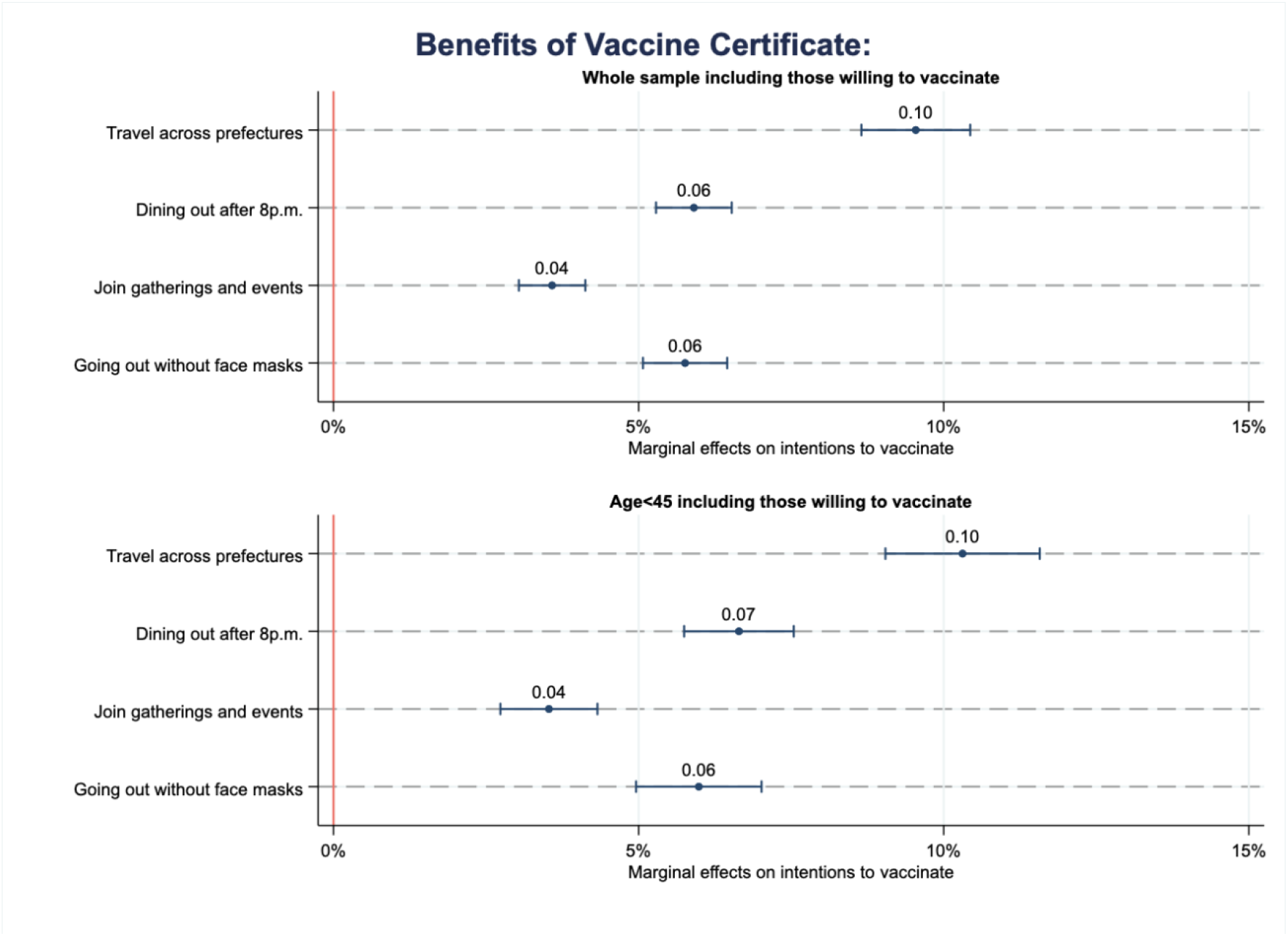
Vaccine passport: Include those willing to vaccinate. Note: Estimates among individuals who have not vaccinated yet, including ‘Unwilling,’ ‘Undecided,’ and ‘Willing’ (n= 3,171 for all age groups and n= 1,644 for those aged 45 or younger); Adjusted for age, gender, co-resident family members, occupation, education, income, health status, statistical literacy, time preference, risk preference, fear of COVID-19, residential area, and vaccine attributes with the population weight for each age group by region; Values and markers are marginal effects with bars representing 95% confidence intervals estimated by robust standard errors; Full results are available upon request.

**Appendix Figure A-8.**
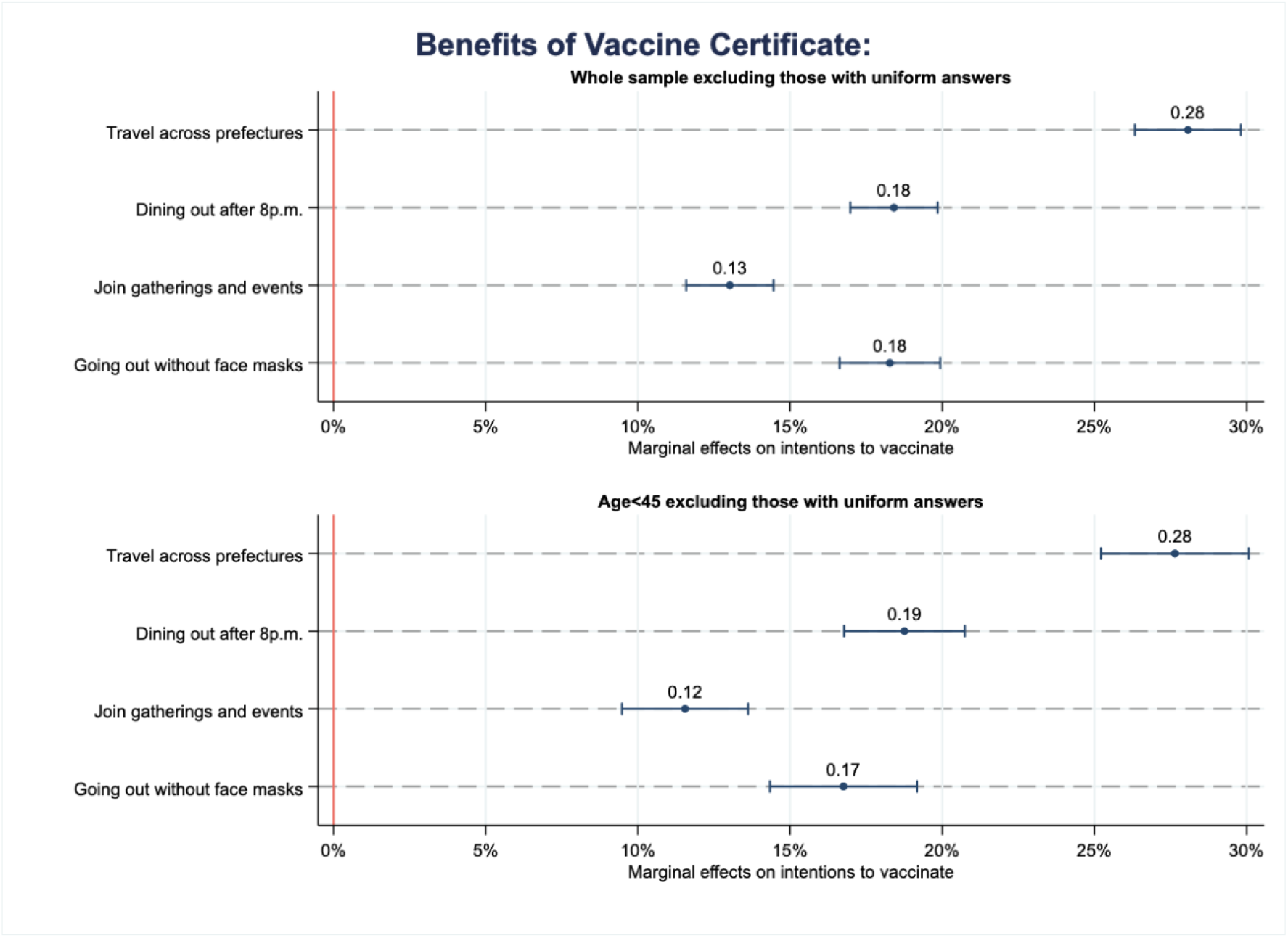
Vaccine passport: Exclude uniform individuals. Note: Estimates among individuals who were undecided and unwilling to vaccinate, excluding those who provided uniform answers regarding their vaccination intentions regardless of available options (n= 1,531 for all age groups and n= 754 for those aged 45 or younger); Adjusted for age, gender, co-resident family members, occupation, education, income, health status, statistical literacy, time preference, risk preference, fear of COVID-19, residential area, and vaccine attributes with the population weight for each age group by region; Values and markers are marginal effects with bars representing 95% confidence intervals estimated by robust standard errors; Full results are available upon request.

**Appendix Figure A-9.**
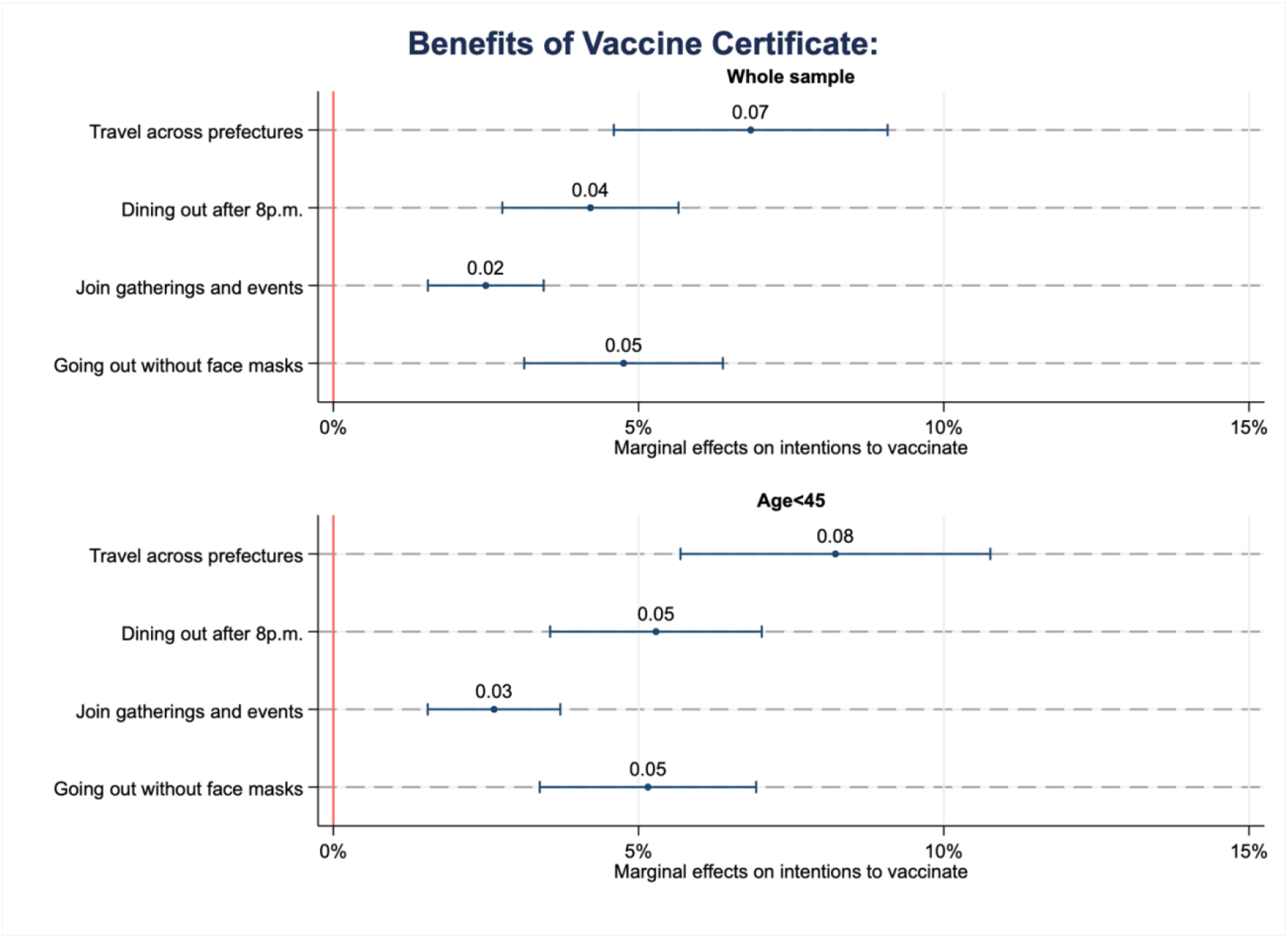
Vaccine passport: Estimates by multilevel mixed-effects logistic regression. Note: Estimated by multilevel mixed-effects logistic regression with random intercepts by individuals among those who were undecided and unwilling to vaccinate (n= 1,518 for all age groups and n= 884 for those aged 45 or younger); Adjusted for age, gender, co-resident family members, occupation, education, income, health status, statistical literacy, time preference, risk preference, fear of COVID-19, residential area, and vaccine attributes with the population weight for each age group by region; Values and markers are marginal effects with bars representing 95% confidence intervals estimated by robust standard errors; Full results are available upon request.

